# Repeated SARS-CoV-2 Introductions with Limited Local Establishment in Bangladesh under Genomic Surveillance, 2020–2025

**DOI:** 10.64898/2026.09.14.26362721

**Authors:** Sabik Khair, Yeasir Karim, Mst. Noorjahan Begum

## Abstract

Genomic surveillance has shown how SARS-CoV-2 lineage composition emerges in densely sequenced, high-income settings. Whether reconstruction is possible where sequencing is increasingly sparse, as in most low- and middle-income countries like Bangladesh, remains untested. To address this, genomes were collected in Bangladesh between March 2020 and July 2025 against a globally distributed background to infer a time-calibrated phylogeny and delineate Bangladeshi transmission lineages. Source attribution was tested against two independent null models that hold background sampling constant. Substitutions enriched in Bangladeshi genomes, relative to a comparator matched on lineage and collection month, were classified as imported or locally arising according to whether they were present at the root of their host lineage. Lineages were inferred at 361 introduction events, of which 51.8% left a single sampled genome with no detected onward transmission. India and the Gulf countries were jointly dominant as source regions, exceeding a rarefied null model by 10.8 and 8.1 percentage points, while East Asia, South-East Asia and Rest-of-World fell significantly below it. Eighteen enriched substitutions, clustered by shared carriers into eleven independent signals, generated 171 substitution-lineage occurrences, of which 53 arose after their host lineage had entered Bangladesh. Spike G446V arose locally in 17 of 19 host lineages and reached 267 of 291 genomes (91.8%) of an imported Delta lineage, indicating that a locally arising substitution could achieve clade expansion. Whether import dependence declined over time could not be evaluated, as the statistic is confounded with sequencing effort. Importation determined which lineages circulated in Bangladesh, while local evolution determined what happened within them. Local substitutions transmitted readily but rarely reached lineage-wide establishment because their host lineages remained under genomic observation for only a short interval.

## 1 Introduction

Genomic epidemiology has established that national SARS-CoV-2 epidemics were not closed systems. Where sampling was dense enough to permit lineage reconstruction, the observed viral diversity largely reflected repeated introductions rather than local emergence. The first wave in the United Kingdom was seeded by more than a thousand independently detectable importations, most of which left few or no sampled descendants, and the largest transmission lineages were disproportionately those introduced before travel restrictions took effect (Du Plessis et al., 2021). Comparable analyses across Europe reached the same conclusion by different routes, the summer 2020 resurgence was driven by reintroduction rather than by persistence of locally circulating lineages (Lemey et al., 2021), the 20E/EU1 lineage spread across the continent on travel rather than by transmission advantage (Hodcroft et al., 2021), and the earliest European and North American clusters were traceable to a small number of distinct introduction events (Nadeau et al., 2021; Worobey et al., 2020). In settings that pursued elimination, the same framing yielded similar results, introductions were numerous but almost none established (Geoghegan et al., 2020; Gu et al., 2022).

The general finding is that the lineage composition of a country is set from outside, and that internal dynamics act as a filter on what arrives rather than as a source of what circulates (Du Plessis et al., 2021; Lemey et al., 2021). Whether this holds where sampling is sparse, intermittent, and geographically concentrated is a separate question, and it matters for two reasons. The first is methodological, where the statistics that carry the conclusion that the number of introductions, the fraction of lineages leaving no sampled descendant, and the median persistence of a transmission lineage depend on the sequencing coverage. A result obtained at high coverage does not transfer to low coverage without argument. The sensitivity of phylodynamic and phylogeographic inference to sampling design is well documented as the reconstruction quality degrades predictably with sampling intensity and strategy (Hall et al., 2016), discrete trait phylogeographic models are biased by unequal sampling across demes because sampling frequency enters the likelihood as though it were migration (De Maio et al., 2015; Liu et al., 2022), and correcting for it requires either explicit modelling of the sampling process (Müller et al., 2018) or the incorporation of external information such as individual travel history (Lemey et al., 2020). These are not abstract concerns in a setting where one country contributes a large share of the tips on a tree. Inferred importation counts have been shown to depend on sampling strategy even under systematic high coverage surveillance (Goliaei et al., 2024). The difficulty takes a sharper form for any rate whose denominator is itself inferred, which to our knowledge has not been examined directly. Importation measured against the amount of transmission already resident requires an estimate of that resident transmission, and in a country whose sequencing effort falls significantly, the estimate falls with it.

The second reason is substantive, as the countries where the question remains unanswered have the largest populations and the densest travel and labor migration links, and are therefore the settings where importation pressure is expected to be highest. Genomic surveillance across Africa has shown both what is achievable and what is lost when sequencing is uneven, as regional lineage dynamics were reconstructed over continents, but with the explicit caution that coverage varied by more than two orders of magnitude between countries (Tegally et al., 2022; Wilkinson et al., 2021). South Asia, which contains roughly a quarter of the world’s population, has received less comparable attention, and we are not aware of a published reconstruction of introduction dynamics spanning the full pandemic for any country in the region (Gustani-Buss et al., 2025).

Bangladesh is such a setting with a population of approximately 170 million, one of the highest densities in the world; shares a 4,000 kilometer land border with India; and maintains one of the world’s largest overseas labor populations, concentrated in the Gulf states (Ahsan et al., 2020; Rahman et al., 2022). It sequenced sufficiently to permit country-scale phylodynamic reconstruction during 2020 and 2021, then sequenced progressively less. Absolute genome output fell from 129 genomes in October 2022 to five in November 2022 and did not recover. 7,854 of the 8,270 deposited genomes (95.0%) were collected before 2023; median monthly output fell from 179 to 4 across that boundary, and 8 of the 38 subsequent months contributed no genomes at all. Reported cases fell in parallel, so sequencing coverage per reported case did not fall but rose, from 0.38 genomes per 100 reported cases in 2021 to 2.61 in 2023 and 2.66 in 2024 (DGHS COVID-19 Dashboard for Bangladesh) and genome counts from GISAID EpiCoV (Shu & McCauley, 2017). The constraint on later-period inference is therefore the absolute number of genomes available to resolve transmission lineages, not the fraction of cases they represent, a distinction that matters because a rate whose denominator is inferred from sampled genomes tracks the former and not the latter. This trajectory is shared by many low- and middle-income countries and is one that any analysis spanning the full pandemic must confront rather than ignore. Published work on SARS-CoV-2 in Bangladesh has largely consisted of descriptive lineage surveys, single-wave analyses, and mutation catalogues (Mohammad Mahmud et al., 2025; Parvin et al., 2021; Sayeed et al., 2022). Although Carnegie et al. studied early transmission dynamics of COVID-19 waves in Bangladesh (Carnegie et al., 2024), no reconstruction of introduction dynamics across 2020–2025 has been reported, and no analysis has asked whether the substitutions common in Bangladeshi genomes arose there.

If importation determined which lineages circulated, then substitutions enriched in Bangladeshi genomes may simply be the substitutions of the lineages that happened to arrive, and their apparent local origin would be an artefact of composition, the same confounding that makes a naive comparison of variant frequencies between countries uninformative. Conversely, a catalogue of Bangladesh-enriched substitutions, without reconstructing their host lineages, cannot distinguish local evolution from a founder effect.

Four processes are easily conflated in a setting like this one, and the analysis was built to separate them. Introduction is the movement of a lineage into Bangladesh. Local transmission is onward spread within the country once a lineage has arrived. Local diversification is the appearance of substitutions on Bangladeshi branches of the tree. Local establishment is a locally arising substitution reaching a defined frequency within its host lineage. Evidence bearing on one does not transfer to the others, and each is reported separately below.

This study aimed to determine whether repeated importation shaped the lineage composition of SARS-CoV-2 in Bangladesh, and whether any substitution arising locally within an imported lineage went on to establish. It found that lineage introductions were frequent and mostly transient, that India and the Gulf countries supplied disproportionately more of them than sampling alone would produce, and that roughly a third of enriched-substitution occurrences arose after their host lineage entered the country. Locally arising substitutions transmitted readily but rarely reached lineage-wide frequency, with one exception of Spike G446V arose independently in most lineages carrying it and swept an imported Delta lineage, indicating that locally arising substitutions can achieve clade-wide expansion. Importation determined which lineages circulated in Bangladesh, and local evolution determined what happened within them.

## 2 Methods

### 2.1 Genomic data, quality control, and the global background

All SARS-CoV-2 genomes with a Bangladeshi collection location deposited in GISAID EpiCoV (Shu & McCauley, 2017) between March 2020 and July 2025 were retrieved on 10^th^ April 2026, yielding 8,270 sequences. Clade, Pango lineage, and per-genome quality metrics were assigned with Nextclade v3.23.0 (Aksamentov et al., 2021), which was used because it produces clade assignment, mutation calling, and quality control from a single reference sequence (MN908947), so that the substitutions later tested for enrichment and the quality metrics used to filter are derived from the same alignment. Genomes were retained for phylogenetic analysis when Nextclade returned an overall quality call of “good” or “mediocre” and genome coverage of at least 90%, and were further subsampled to 4598 sequences grouped by Collection month and lineage, apart from the sequences from 2024 and 2025, whose sequences were retained even after a “bad” score from Nextclade with coverage >95% and the frameshifts were excluded. The recombinant variants were retained because pruning tips would desynchronize the maximum clade credibility (MCC) tree from the posterior sample.

The background genomes were retrieved from GISAID EpiCoV and assigned to one of seven source regions by country: India, the Gulf (Saudi Arabia, Qatar, United Arab Emirates, Kuwait, Bahrain, Oman), Europe, South-East Asia (Myanmar, Thailand, Singapore, Malaysia, Indonesia, Laos, Cambodia, Vietnam), East Asia (China, Japan, South Korea), Africa, and Rest-of-World (Table S1a). The selection was stratified by region and calendar month using the GISAID EpiCoV “Subsample” grouping by “Collection date by month” and “Lineages”, with genome coverage >95% for most subsampling (Few sequences with >90% coverage were included to maintain temporal homogeneity). The background is deliberately not proportional to global deposition. Regions with substantial travel and labor migration links to Bangladesh are represented more heavily than their share of GISAID submissions would give (Ahsan et al., 2020; Rahman, 2022).

### 2.2. Alignment, molecular dating, phylogenetic and statistical inferences

The SARS-CoV-2 complete genomes were aligned with MAFFT v7.511 (Katoh & Standley, 2013) and a maximum-likelihood topology was estimated in FastTree v2.1.10 (Price et al., 2009) under the GTR+Γ model with other parameters at default, and the temporal signal was assessed by root-to-tip regression in TempEst v1.5.3 (Rambaut et al., 2016), which provided significant divergence accumulation with sampling date.

Divergence times were estimated in BEAST v1.10.5pre with the ThorneyBEAST package v0.1.2 (Du Plessis et al., 2021; Suchard et al., 2018). ThorneyBEAST was used because it assumes a fixed tree topology and re-estimates only node heights, which makes Bayesian dating of a 10,995-tip alignment computationally feasible. However, no substitution-model parameters are sampled, so this analysis yields no independent estimate of the substitution process. A strict molecular clock was applied with the rate fixed at 8.0 × 10⁻⁴ substitutions site⁻¹ year⁻¹, and a tree prior SkyGrid coalescent model was used (Gill et al., 2013), chosen as a flexible non-parametric prior. Three independent chains were run for 200 million chains, sampling every 10⁵ states, under seeds 42, 123 and 999 and pooled with LogCombiner, and a maximum clade credibility tree was summarised with TreeAnnotator using the common ancestor, with 10% burn-in states; 999 trees were drawn at even intervals from the pooled posterior for all replicate analyses.

Quantities estimated across the posterior tree sample are summarized by the median and a 95% highest posterior density interval, computed as the narrowest interval containing 95% of the sampled values; this is distinguished throughout from an equal-tailed percentile interval, which for the skewed posteriors here gives different bounds. Intervals on quantities that are not posterior samples are identified as such at each use. Where a permutation test reaches the smallest p-value its design can attain and effect sizes are interpreted in place of significance. Analyses were implemented in Python 3.12 (pandas 3.0.3, NumPy 2.4.6, SciPy 1.15.3) and R 4.3.1 (ggplot2 4.0, patchwork 1.3.2, dplyr 1.2.1)

### 2.3 Ancestral state reconstruction and transmission-lineage delineation

Ancestral geographic states were reconstructed with PastML v1.9.50 (Ishikawa et al., 2019) under the marginal posterior probabilities approximation (MPPA) with an F81 substitution model. Two traits were reconstructed as a Bangladesh/non-Bangladesh trait used to delineate lineages, and an eight-state trait including Bangladesh and the seven background regions, used for source attribution, with the same sequences in separate PastML runs.

A Bangladeshi transmission lineage (BDTL) is an internal node reconstructed as Bangladesh whose parent is reconstructed as non-Bangladesh, together with all its descendants. This strict criterion counts every geographic transition as an introduction. A relaxed criterion additionally requiring a Bangladeshi grandparent was applied in parallel and is reported as a lower bound, since it merges introductions that are nested within one another. Delineation was applied independently to the MCC tree and to each of the 999 posterior trees. Lineages are matched across trees by the hash of their exact tip set, not by clade position, so that the comparison is independent of topology. Posterior support is the fraction of the 999 trees in which the identical tip set is delineated as a BDTL.

Persistence is the interval from a lineage-inferred TMRCA to its most recent sampled genome, and detection lag is the interval from the TMRCA to its first sampled genome. Persistence contains the detection lag by construction. Persistence statistics are computed on lineages recovered at support above 0.5, because exact tip-set recovery is easier for small lineages, that set is enriched for them, and the resulting medians are reported as lower bounds.

### 2.4 Source attribution and control for sampling imbalance

Each introduction was attributed to a source region from the probability-weighted marginal reconstruction of its parental node under the eight-state character. Hard maximum *a posteriori* assignment was computed as a sensitivity analysis. Attribution counts each introduction once, irrespective of how many genomes descend from it.

Raw attribution shares are inflated by unequal background sampling, because a region contributing more tips is reconstructed at more internal nodes. Two independent controls were therefore applied. In every control arm, lineage boundaries were held fixed at the binary reconstruction, so all arms delineate an identical set of introductions and differ only in the deme labels available for attribution. This isolates the attribution step from the delineation step, which would otherwise vary together.

Randomized-trait null (10 replicates) were reconstructed with region labels permuted across background tips, destroying geographic signal while preserving tip abundance exactly. A reconstruction that carries genuine geographic information should perform better than this model; one that merely tracks abundance will not. Subsequently, rarefied null models (10 replicates) were reconstructed where every background region was subsampled to 450 tips and the same label permutation applied. Because abundance is equalized by construction, any departure of this null from the uniform expectation of 14.3% measures bias attributable to topology or branch length structure rather than to sampling distribution.

Rarefied and rarefied-null model replicates are paired by construction; replicate *i* of the null estimate is a label permutation of replicate *i* of the rarefied trait file and shares its tip set. Excess attribution was therefore tested by an exact paired sign-flip permutation test enumerating all 2¹⁰ = 1,024 sign assignments. Effect sizes are reported as the mean paired difference divided by its standard deviation, and are interpreted separately from significance.

### 2.5 Importation per unit resident lineage-time

λ(t) is the number of introductions arriving in a quarterly slice divided by the total Bangladeshi lineage-time at risk in that slice, computed independently on each of the 999 posterior trees. It expresses importation relative to the amount of transmission already resident.

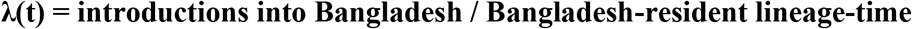

Each branch whose child node is reconstructed as Bangladesh contributes its overlap with each slice. The stem branch subtending a lineage root is excluded as geographic transition occurred at an unknown point along that branch. Therefore, counting the whole of it inflates the denominator and yields negative residence ages for genomes sampled early in a lineage. Exclusion is the conservative choice, because it removes time that is disproportionately imported recently, working against an import-driven conclusion. Alternative tests with half the stem and the whole stem were implemented as sensitivity analyses. The primary window ends at 2024.0, the point from which national genomic surveillance had collapsed and annual introduction totals cease to be comparable with earlier periods. This boundary was fixed by the surveillance record rather than chosen to maximise the estimate; an earlier boundary at 2023.5 yields a larger slope. No minimum-lineage-time criterion is applied to slices inside the window. Windows ending at 2023.0, 2023.5 and 2024.5, the untruncated series, and a restriction to slices holding at least five lineage-years are reported as sensitivity analyses.

Resident lineage-time is not observed but estimated from sampled genomes, so it falls when sequencing falls, whether or not transmission does. The relationship was characterized by regression of log lineage-time on log genomes sequenced per slice. λ was then adjusted by adding genomes sequenced per slice as a covariate to a linear model of λ on slice midpoint and refitting on every posterior tree, taking the coefficient on the midpoint. The covariate entered on the raw scale where sequencing counts were taken from the national surveillance record aggregated to the same quarterly boundaries. Because the confounding relationship was characterized on the logarithmic scale while the adjustment is additive, the analysis was repeated with the covariate entered logarithmically, and both are reported. Slices within the window differ by two orders of magnitude in the lineage-time they carry. The unadjusted and adjusted fits were therefore repeated with the single slice carrying the highest λ inside the window excluded.

In contrast, P_local, the proportion of Bangladeshi lineage-time attributable to locally sustained transmission, was computed in three variants distinguished by the residence age at which lineage-time is counted as local (14, 30 and 60 days), together with the mean residence age. Trends were estimated by fitting a slope across slices on every posterior tree and summarizing the resulting distribution, and were evaluated across five thresholds for slice inclusion.

### 2.6 Substitution enrichment, origin attribution and artefact screening

Substitutions observed in Bangladeshi genomes were tested for enrichment matching against lineage and month global comparator. Genomes in both arms were assigned to strata defined by Pango lineage and collection month, and within each stratum a 2 × 2 table cross-classified carrier status by arm. Evidence was pooled across strata with the **Cochran–Mantel–Haenszel (CMH) test** (Ghoshal & Chen, 2025; Mantel & Haenszel, 1959), which was used because it holds lineage and calendar time constant by design, an unstratified comparison of frequencies between countries measures which lineages circulated where. Significance used the CMH χ² with Benjamini–Hochberg correction (Benjamini & Hochberg, 1995). Carrier sets overlap substantially, so the tests are not independent.

Each enriched substitution was encoded as a binary presence or absence of mutations and reconstructed on the MCC tree in a PastML run separate from the geographic characters, under the same MPPA/F81 settings. Reconstructions were screened for convergence. A mutation whose fitted branch-scaling factor sits at the upper bound of the optimizer has not converged, at that rate the F81 transition probabilities approach the stationary state frequencies irrespective of branch length, so each node is assigned the more frequent state essentially independently of its neighbors. Such a reconstruction returns no unresolved nodes and drives every root posterior toward zero, producing a local-origin call by construction. Non-converged characters were excluded.

An **occurrence** is a substitution-lineage pair. It is classified as **imported** when the substitution is present at the root of its host lineage and **locally arising** when absent, on a threshold of 0.5 applied to the root marginal probability. Threshold sensitivity was assessed by sweeping the cut across the interval separating the two classes. Two derived quantities are defined on occurrences. **Onward transmission**, where the substitution is carried by at least two genomes of its host lineage and is reported for locally arising occurrences only. It is not compared between origin classes as an imported substitution sits at its lineage root by construction and, absent reversion, is carried by all descendants, so its carrier fraction is a property of the classification rather than a measure of spread. **Establishment,** where the mutation is locally arising, carried by at least ten genomes, and reaching at least half the host lineage.

Three independent screens were applied to the candidate set for understanding the artefacts. Codon positions were converted to Wuhan-Hu-1 nucleotide coordinates, using the convention in which ORF1b residue numbering begins at the −1 ribosomal frameshift site, and intersected with the community problematic-sites resource (De Maio et al., 2024). Moreover, substitutions adjacent to an indel were flagged in the enrichment step, because alignment ambiguity at indel margins produces spurious substitution calls. Furthermore, a substitution common in one country and rare globally may reflect sequencing bias in a laboratory that contributed many genomes from that country. Submitting-laboratory records were obtained for all Bangladeshi tips. For each substitution, carrier status was permuted within strata (1,000 replicates, seed 42) and the observed top-laboratory share among carriers was compared with the permuted null values, with Benjamini–Hochberg correction (Benjamini & Hochberg, 1995).

The **primary stratification was Pango lineage × collection month × district**. District is included because a substitution that spread within one district would be laboratory-concentrated for epidemiological rather than technical reasons, where laboratories sample geographically unevenly. Stratification by Pango lineage × month alone is reported as a sensitivity analysis, and the two are interpreted together using the proportion of carriers sitting in a stratum that admits more than one permutation where a substitution losing significance under the finer design while retaining permutability has had a confound removed, whereas one losing both has lost power.

Enriched substitutions carried on a shared genomic background are not independent observations. The substitutions were clustered by single linkage on the Jaccard index of their Bangladeshi carrier sets. Single linkage is appropriate because co-inheritance is transitive along a haplotype; two substitutions may share few carriers while both sharing many with a third, and all three still sit on one background. The threshold is a convention rather than an estimate, so results are reported across a sweep (Jaccard 0.30, 0.50, 0.70, 0.95) with 0.50 as primary.

Blocks are not merged in the reconstruction, as each character remains a separate ancestral-state character, and its occurrences remain separate occurrences. What changes is the counting. Claims about how many distinct Bangladesh-enriched changes occurred are made over blocks; claims about the reconstruction are made over characters; and an establishment event involving several members of one block within one lineage is counted once.

## 3 Results

### 3.1 Dataset, reconstruction quality, and temporal signal

Of 8,270 SARS-CoV-2 genomes submitted from Bangladesh between March 2020 and July 2025, 8,232 were human-derived and formed the epidemiological denominator; excluded 38 environmental samples from case-based analyses. Nextclade v3 returned an overall quality call of good for 6,011 genomes, mediocre for 1,137, and bad for 1,080, with four producing no output (Table S2). An additional coverage threshold was imposed at 0.90; the intersection of good or mediocre with ≥0.90 coverage retained 6,985 genomes, which were further subsampled to 4,598 BD sequences grouped by month and lineage for the dated tree.

The time-calibrated phylogeny comprised 10,995 tips, Bangladeshi and globally distributed background (Fig. 1). Background composition was India 1,543 tips (24.1%), Gulf 1,067 (16.7%), Europe 1,023 (16.0%), South-East Asia 968 (15.1%), East Asia 819 (12.8%), Africa 527 (8.2%) and Rest-of-World 450 (7.0%) (Table S1a and Table S1b). The root-to-tip slope exceeds the fixed clock rate used for dating. The regression was used as a temporal-signal diagnostic and not as a rate estimate. It is fitted on a tree spanning five variant sweeps and retaining recombinant genomes, both of which inflate the apparent slope. The prior rate of 8.0 × 10⁻⁴ substitutions site⁻¹ year⁻¹ was retained. Root-to-tip regression supported a clock-like signal (slope 1.273 × 10⁻³ substitutions site⁻¹ year⁻¹, R² = 0.924), and a date-randomization test rejected the null of no temporal structure (p < 0.001). As an external check, the resulting root TMRCA of 2019.886 [2019.827, 2019.949] is concordant with independent estimates of the pandemic origin.

**Figure 1:**
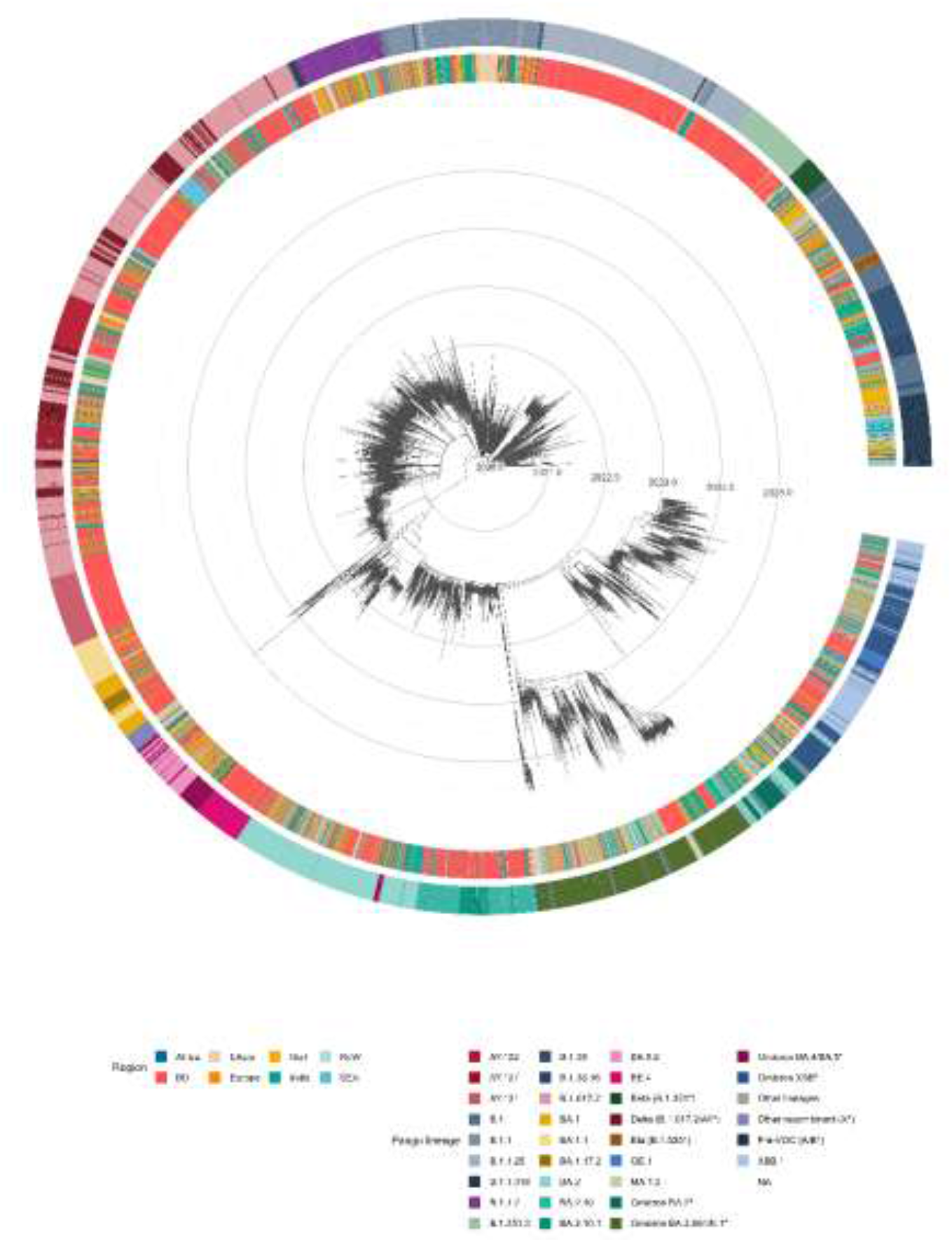
Time-scaled phylogenetic tree of Bangladeshi SARS-CoV-2 strains against a global backbone. Concentric rings mark calendar years; root TMRCA 2019.886, scale bar = 1 year. Inner strip: Regions - Africa, EAsia (East Asia), Gulf countries, BD (Bangladesh), Europe, India, SEA (Southeast Asia), RoW (Rest of the World); Outer strip: Pango lineage. 10995 tips (4598 Bangladesh).

Ancestral state reconstruction (PastML, MPPA/F81) was well resolved (Table S3). The binary Bangladesh vs. non-Bangladesh character left 193 of 10,994 internal nodes unresolved (1.76%), a node is unresolved when the method retains more than one state. Resolution elsewhere on the tree was near-complete: the median maximum marginal probability across nodes was 1.000, and 95.9% of nodes exceeded 0.99. The fitted Bangladesh equilibrium frequency was 0.318, compared with an observed tip frequency of 0.418. In contrast, the seven-region character left 1,116 nodes unresolved (5.08%; scaling 1.615) and placed the root in East Asia with posterior probability 0.9994.

### 3.2 Introductions were frequent, mostly transient, and heavy-tailed in size

Applying the strict criterion, an internal node reconstructed as Bangladesh whose parent is reconstructed as non-Bangladesh, across 999 posterior trees identified 361 independent introductions (95% HPD 305–409; Fig. S1). A relaxed criterion additionally requiring a Bangladeshi grandparent yielded 152 (69–260) and is reported as a lower bound only. The two reconcile exactly: of 395 strict lineages delineated on the MCC tree, 143 are nested within larger Bangladeshi clades, and 252 are top-level, which are also the relaxed count.

The size distribution of Bangladeshi transmission lineages (BDTLs) is heavily skewed (Fig. 2a). Across the posterior, the median per-tree fraction of singleton lineages, one genome descending from one introduction with no sampled onward transmission, is 51.8% (95% interval 49.0–54.3), corresponding to a posterior median of 187 singleton lineages. A further 21.1% (18.9–23.6) contain at least 5 genomes, 13.5% (11.8–15.3) at least 10, 4.1% (3.6–4.7) at least 50, and 2.5% (2.0–3.2) at least 100. On the MCC tree specifically, these correspond to 205 singletons and 86, 53, 14, and 8 lineages of 395. The MCC yields slightly more lineages but slightly fewer large ones than the typical posterior tree (395 against 361; 8 against 9 of ≥100 genomes), because a consensus summary splits large clusters marginally more often than an individual tree does. The complementary cumulative distribution has an exponent of −1.08 (R² = 0.72), stable between −1.08 and −1.16 for minimum sizes of 1 to 10; this was reported descriptively and does not claim a scale-free distribution. The posterior median of the per-tree largest lineage is 843 (841–843); the pooled distribution across all 999 trees extends to 1,068 genomes (Fig. 2a).

**Figure 2:**
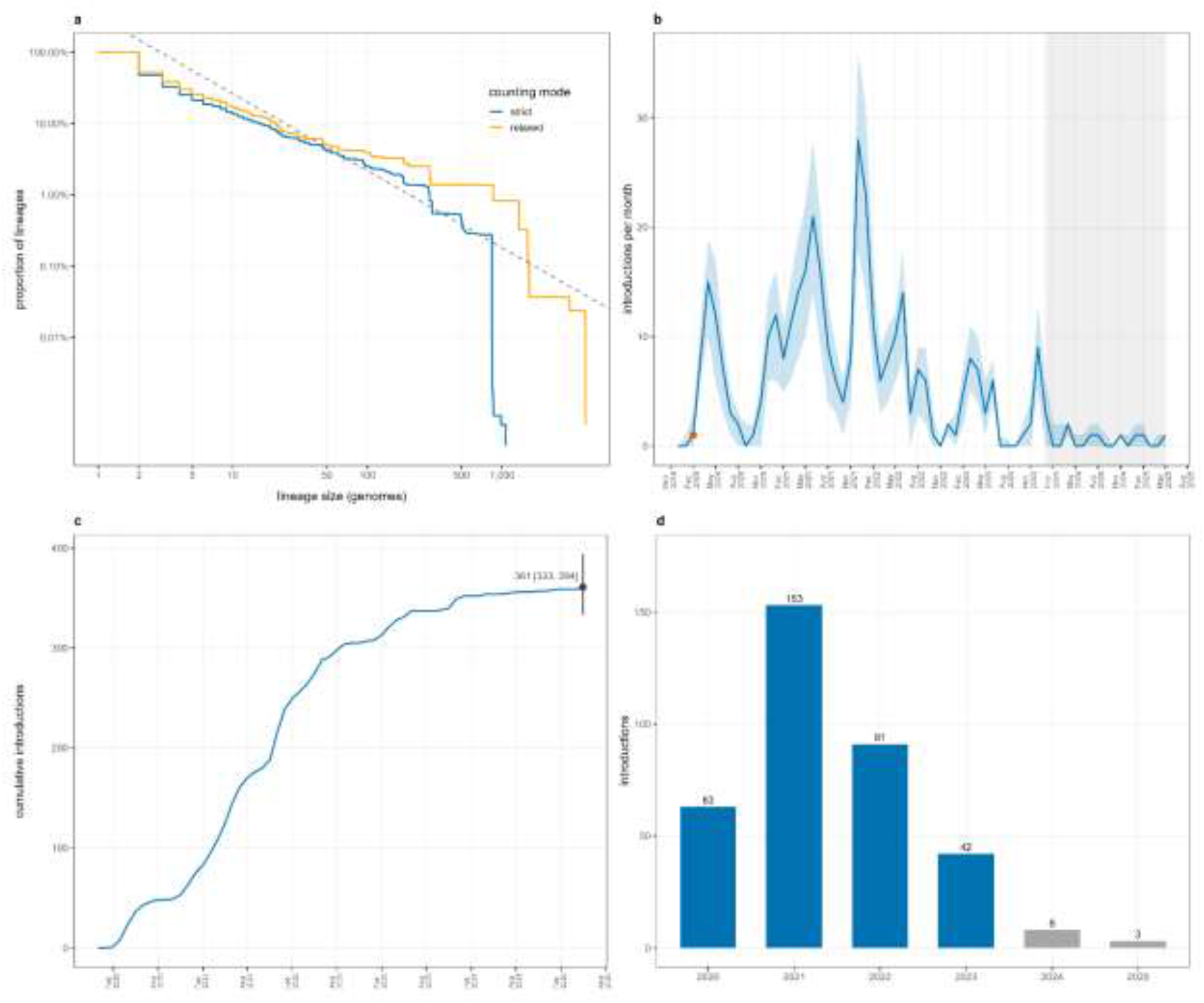
Introduction counts of SARS-CoV-2 lineages in Bangladesh. (a) Bangladeshi transmission lineage sizes are heavy-tailed. Pooled over 999 posterior trees; pooled maximum of 1068 genomes. The posterior median of the per-tree largest lineage is 843 [841, 843]. 51.8% singletons, CCDF exponent: −1.08 (R^2^ = 0.72), stable at −1.08 to −1.16 for x_min 1-10. (b) Inferred introductions into Bangladesh. Posterior median and 95% HPD across 999 trees; series spans Dec 2019 to May 25. The post-2023 decline coincides with the collapse of national genomic surveillance and cannot be interpreted as reduced importation; the shaded interval in marks this period. (c) Cumulative introductions. Monthly medians sum to 360; the posterior median total is 361. Medians are not additive, and Bounds are the posterior HPD of the total, not a cumulative sum of monthly HPDs. (d) Yearly total introductions. 2020 and 2025 are partial years. Grey bars fall inside the surveillance collapse and are not comparable.

Persistence, defined as the interval from an inferred TMRCA to its most recent sampled genome, scales steeply with lineage size, and a single median would misrepresent it. This was computed for the 312 lineages whose exact tip set recurs in the posterior at support above 0.5, of which 179 are singletons, and 133 contain more than one genome. Among those 133, the median is 62 days, rising to 163 days at ≥ 10 genomes, 273 days at ≥ 50, and 321 days at ≥ 100 (Fig. S2). Singletons have zero persistence by construction and are excluded. Because exact tip-set recovery is easier for small lineages, singletons comprise 57.4% of the support-filtered set against 51.8% of the posterior as a whole; this set is enriched for small lineages, and the 62 day median should be read as a lower bound. The corresponding median for the 53 MCC lineages with ≥10 genomes is 177 days.

For the median interval from TMRCA to the first sampled genome, the detection lag was 48 days across the posterior (95% HPD 10–156) and 46 days (IQR 33–63) among the 53 MCC lineages with≥10 genomes. Because persistence is measured from the TMRCA, it includes the detection lag of a lineage with a 46-day lag, and 62 days of persistence was sampled across a 16-day window after roughly six weeks of undetected circulation.

Importation was strongly episodic (Fig. 2b). Introductions peaked at 28 per month (HPD 18–36) in December 2021, coincident with the arrival of Omicron BA.1. Yearly totals were 63 (2020), 153 (2021), 91 (2022), 42 (2023), 8 (2024) and 3 (2025) where 2020 and 2025 are partial years (Fig. 2d). The earliest inferred introduction falls in February 2020, one month before the first laboratory-confirmed case on 8 March 2020 in Bangladesh, and is present in 920 of 999 posterior trees. Sampling began with the first sequenced genome in March 2020; the February inference is cryptic circulation preceding detection.

Posterior support was assessed at the lineage level: 312 strict lineages exceeded support 0.5, 161 exceeded 0.9, and 96 exceeded 0.99 (Fig. S3). Two distinct objects must be distinguished. However, the MCC lineage table (Table S4) contains 53 lineages of ≥10 genomes delineated on the MCC tree, of which 33 are recovered as supported tip sets in the posterior sample, and 20 are not; the posterior support file contains 35 supported lineages of ≥10 genomes. This is because the posterior supported lineages were defined by their descendant tip sets rather than by their position on the MCC tree. Unique tip sets were tracked using hash-based identifiers, allowing lineage membership to be compared across posterior trees despite differences in topology. The unsupported lineages are not uniformly small. Four of the ten largest MCC lineages, BDTL-002 (497 genomes), BDTL-007 (181), BDTL-008 (131), and BDTL-009 (97), were not recovered as supported tip sets in the posterior at all and carry no support value. Thus, although these clusters are delineated on the MCC tree, their exact genome composition was not corroborated across posterior trees. This distinction is important: lack of posterior support for a tip set does not imply that the corresponding cluster is absent, but rather that its membership is uncertain. The restricted lineage-level was therefore inferred to the 33 posterior-supported lineages, and the remaining 20 MCC-defined lineages were reported for completeness. BDTL-001, the largest lineage (843 genomes), received posterior support of 0.868. An independent observation points in the same direction and does not depend on sequencing depth, as the counts above. Among genomes passing quality control, 424 were assigned to Beta (B.1.351 sub-lineages) and 358 to Alpha (B.1.1.7), comparable abundance, but very different internal structure. 311 of the 424 Beta genomes (73.3%) belong to the single sub-lineage B.1.351.3, attributed to an African source at posterior probability 0.975, in a country where Africa is otherwise indistinguishable from the rarefied null model values as a source of introductions (section 3.3).

Together, 361 introductions, over half with no sampled descendants, a median persistence of two months, and variant composition determined by which lineages arrived, importation rather than local diversification determined which lineages circulated.

### 3.3 Introductions came disproportionately from India and the Gulf countries

Attributing each introduction to a source region by its probability-weighted marginal reconstruction, India accounted for 37.8% of introductions (95% HPD 34.6–41.3) and the Gulf for 26.9% (23.9–29.9) as posterior medians across 999 trees, against background shares of 24.1% and 16.7% (Table 3). On the MCC tree alone, the corresponding point estimates are 39.9% and 26.9%. Hard maximum-a-posteriori assignment changes little (42.0% and 27.1%). Attribution counts each introduction once, irrespective of how many genomes descend from it. To address the possible inflation by unequal background sampling, two independent control models were applied (Fig. 3).

**Figure 3:**
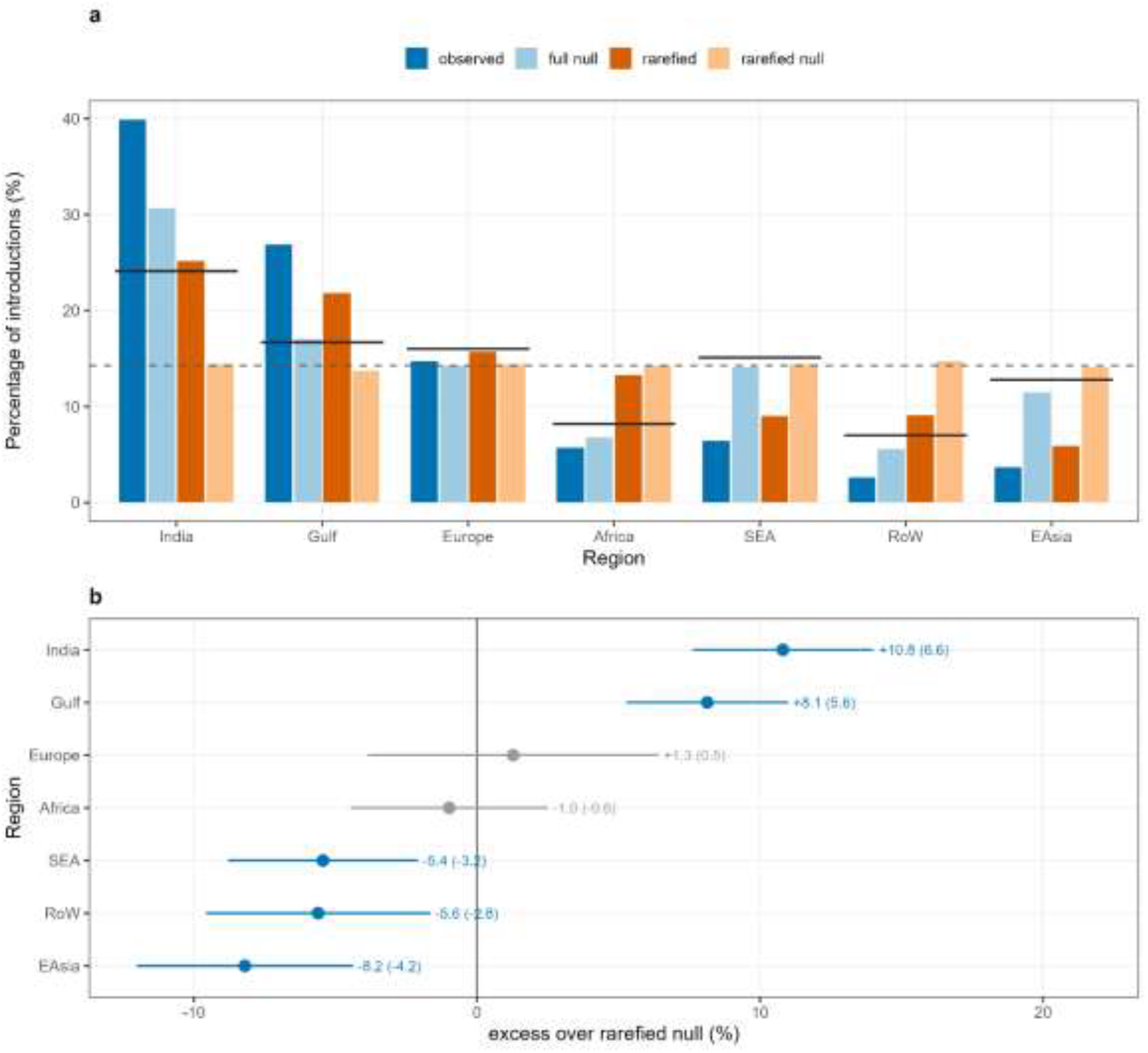
Proportion of lineage attribution from each region. (a) Attribution before and after each control. The horizontal dashed line (black) spans each group = background tip share in the full dataset. Under rarefaction, every null return to uniform, so the distortion in the full analysis is abundance-driven, not topological. (b) Supported sources and deficits. Paired differences ± 1.96 replicate SD. Significance from the exact paired sign-flip test (floor p = 0.002 with 10 pairs, reached by all five significant regions); SD values are effect sizes. India and Gulf differ by ∼1.2 SD and are not ranked.

**Table 1:**
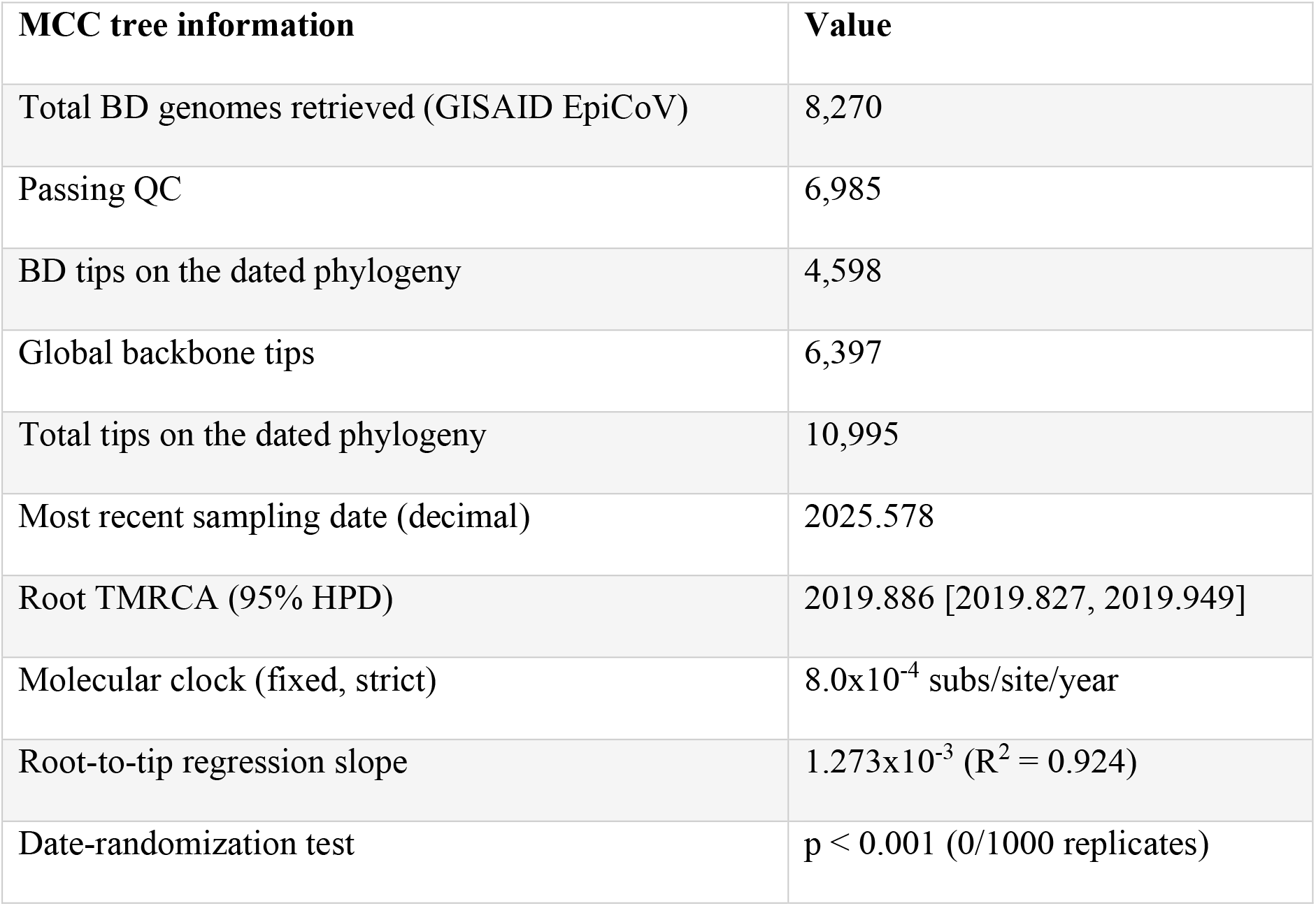
Time-scaled MCC phylogenetic analysis parameters.

**Table 2:**
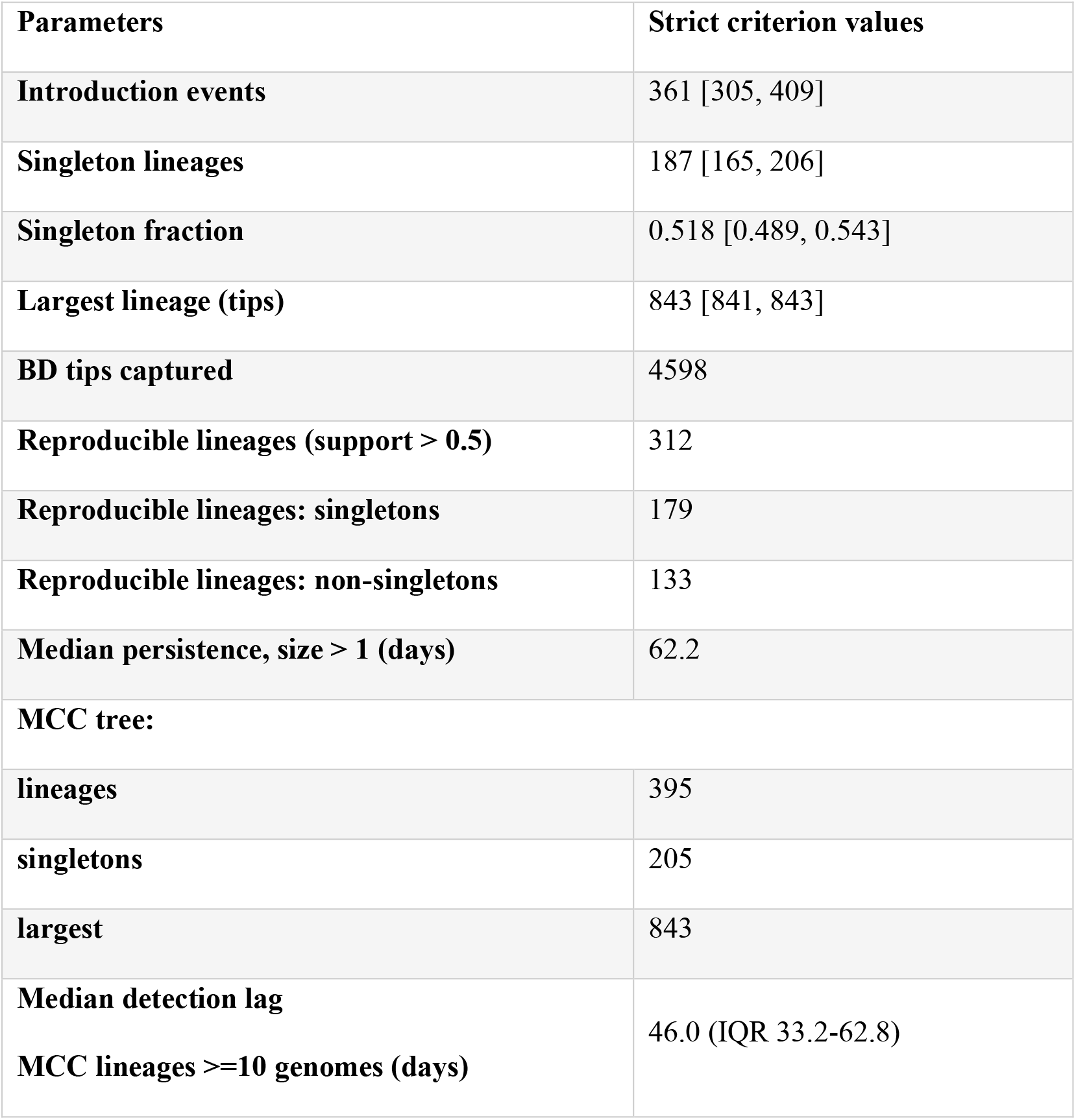
Parameters on strict criterion values of lineage introduction.

**Table 3:** Source attribution of Bangladeshi introductions, before and after two controls for sampling imbalance.

| **Source region** | **Introductions ([95% HPD])** | **Full backbone (%)** | **Rarefied (n=450)** | **Rarefied null model (%)** | **Excess over null model estimates (pp)** | **p** |
| --- | --- | --- | --- | --- | --- | --- |
| **India** | 136.8 [120.7, 157.0] | 37.90 | 25.2 [22.3, 28.3] | 14.4 | 10.8 | 0.002 |
| **Gulf** | 97.7 [81.6, 110.1] | 27.00 | 21.8 [18.5, 24.3] | 13.7 | 8.1 | 0.002 |
| **Europe** | 49.3 [42.3, 57.2] | 13.60 | 15.5 [13.4, 18.6] | 14.2 | 1.3 | 0.15 |
| **SEA** | 29.0 [22.1, 35.3] | 8.00 | 9.0 [7.9, 10.3] | 14.4 | -5.4 | 0.002 |
| **Africa** | 21.6 [16.8, 26.5] | 6.00 | 13.4 [11.8, 15.0] | 14.4 | -1.0 | 0.13 |
| **East Asia** | 14.6 [10.8, 18.2] | 4.00 | 5.6 [4.8, 7.5] | 13.8 | -8.2 | 0.002 |
| **RoW** | 12.5 [9.0, 16.7] | 3.40 | 9.1 [7.0, 11.2] | 14.7 | -5.6 | 0.002 |

First, a randomised-trait null (10 replicates) permuting region labels across background tips. Mean hard-assignment confidence was 0.834 against a null model of 0.630 ± 0.014, the median 0.928 against 0.614 ± 0.026, and the fraction assigned at posterior >0.9 was 0.549 against 0.148 ± 0.021; 0 of 10 replicates reached the observed value. The null estimate attribution profile tracked background tip abundance (r = 0.977), confirming it behaves as expected (Table S5).

Second, rarefaction of every background region to 450 tips (10 replicates; pairwise Jaccard similarity 0.703–0.711) was combined with the same permutation. This rarefied null estimate isolates bias with abundance removed, and its behavior is informative in itself, as every rarefied null estimate of the regions falls within 0.6 percentage points of the uniform 14.3% expectation (Fig. 3a). Under equalized tip abundance, the null model returns to within 0.6 percentage points of uniform for every region, so the distortion observed in the uncontrolled analysis is attributable to sampling abundance rather than to topology or branch-length structure. This establishes that the reconstruction procedure is not distorted by the tree itself; it does not establish that the real-world attribution is unbiased, since rarefaction models equalizes tip counts but not epidemic size, sampling period, within-region diversity, or the unsampled countries absent from the background altogether. Removing the reference genome from the rarefaction pool displaces no region by more than 1.2 replicate standard deviations (Fig. S4).

Rarefied and rarefied-null model replicates are paired by construction: replicate *i* of the null is a label permutation of replicate *i* of the rarefied trait file and shares its tip set. Both arms delineated exactly 395 lineages in every replicate, confirming that boundaries were held fixed. Excess attribution was therefore tested by exact paired sign-flip permutation over all 2¹⁰ = 1024 sign assignments; the two-sided floor is 0.00195.

Two regions are elevated against this fully controlled null, India by 10.8 and the Gulf by 8.1 percentage points (pp) in all 10 replicates. Three regions fall significantly below, which are East Asia (−8.2 pp), RoW (−5.6 pp), and South-East Asia (−5.4 pp) (0/10 replicates). Europe (8/10 replicates) and Africa (3/10 replicates) are indistinguishable from the null model (Fig. 3b).

All five significant results sit at the permutation floor, so p does not discriminate among them; effect size does. Under this primary analysis, India excess exceeds the Gulf by 2.66 pp, favoring India in 9 of 10 replicates. The two were reported as jointly dominant rather than ranked, because that ordering is not stable: under the uncontrolled full-data null model, the Gulf exceeds India (9.9% against 9.2%).

The combined deficit of East Asia, RoW and South-East Asia (−19.2 pp) almost mirrors India– Gulf (18.9 pp), consistent with a greater contribution from regions linked to Bangladesh by major labour-migration and air-travel routes than from geographically proximate ones (Sayeed et al., 2022). East Asia deficit is notable given that the root is reconstructed there at posterior 0.9994, indicating the initial corridor closed and did not reopen.

Africa is not supported as a source corridor. Its excess over the rarefied null model is −1.0 percentage (3 of 10 replicates, p = 0.13), and an earlier reading of Africa rise under rarefaction as evidence of suppressed signal was measured against the wrong reference. The high-confidence B.1.351.3 attribution is a single, independently corroborated introduction event, not a general corridor. That one event founded a large lineage, and only tested for the number of introductions arriving from a region, not the size of the lineages.

Source attribution is not reported per lineage (Table S4) for two reasons. Thirteen of the 53 MCC lineages of ≥10 genomes have source posteriors below 0.75 and four below 0.5 (BDTL-022, Gulf 0.42; BDTL-029, Europe 0.49; BDTL-036, Gulf 0.38; BDTL-053, India 0.46); these include four of the five largest lineages, whose deep and mixed backbone ancestry makes them the least confidently attributed rather than the most. More importantly, the modal source call changes under rarefaction for 16 of the 53 (30%), and seven of those move to Africa or Rest-of-World, the two demes that are null in the aggregate analysis. Because rarefaction is the primary control, a lineage-level call that survives only on the unrarefied backbone cannot be reported.

Table S4 therefore gives each lineage source posterior and a rarefaction-stability flag but not a named region. The aggregate attribution is unaffected as it is computed over all 395 introductions with boundaries held fixed, and individual lineage instability is the sampling noise that the paired design averages over. One low-confidence call is also biologically implausible and illustrates the point that BDTL-005 is a well-supported Delta clade (support 0.754) attributed to Africa (0.689).

### 3.4 Importation per unit resident lineage-time is confounded with sequencing effort

If Bangladesh had progressively acquired autonomous transmission, the rate of new introductions per unit of resident Bangladeshi lineage-time, λ(t), should have fallen, which did not. The estimated trend is positive, but surveillance effort confounds it in a way these data cannot resolve, so the direction was reported and no conclusion was drawn about whether import dependence changed.

λ(t) is the number of introductions arriving in a quarterly time slice divided by the total Bangladeshi lineage-time at risk in that slice. From 2020.1 to 2024.0, the unadjusted slope is 0.459 per year (95% HPD 0.205 to 0.713). The mechanism is a contracting denominator rather than accelerating importation, as introductions per quarter fell from a peak near 60 in 2021 to fewer than 10 after 2023, while resident lineage-time fell from 112 lineage-years in mid-2020 to 1.4-7.2 across 2023-2025 (Fig. 4a). Five slices inside the primary window carry fewer than five lineage-years and are shown separately, because λ is unstable where the denominator is that thin.

**Figure 4:**
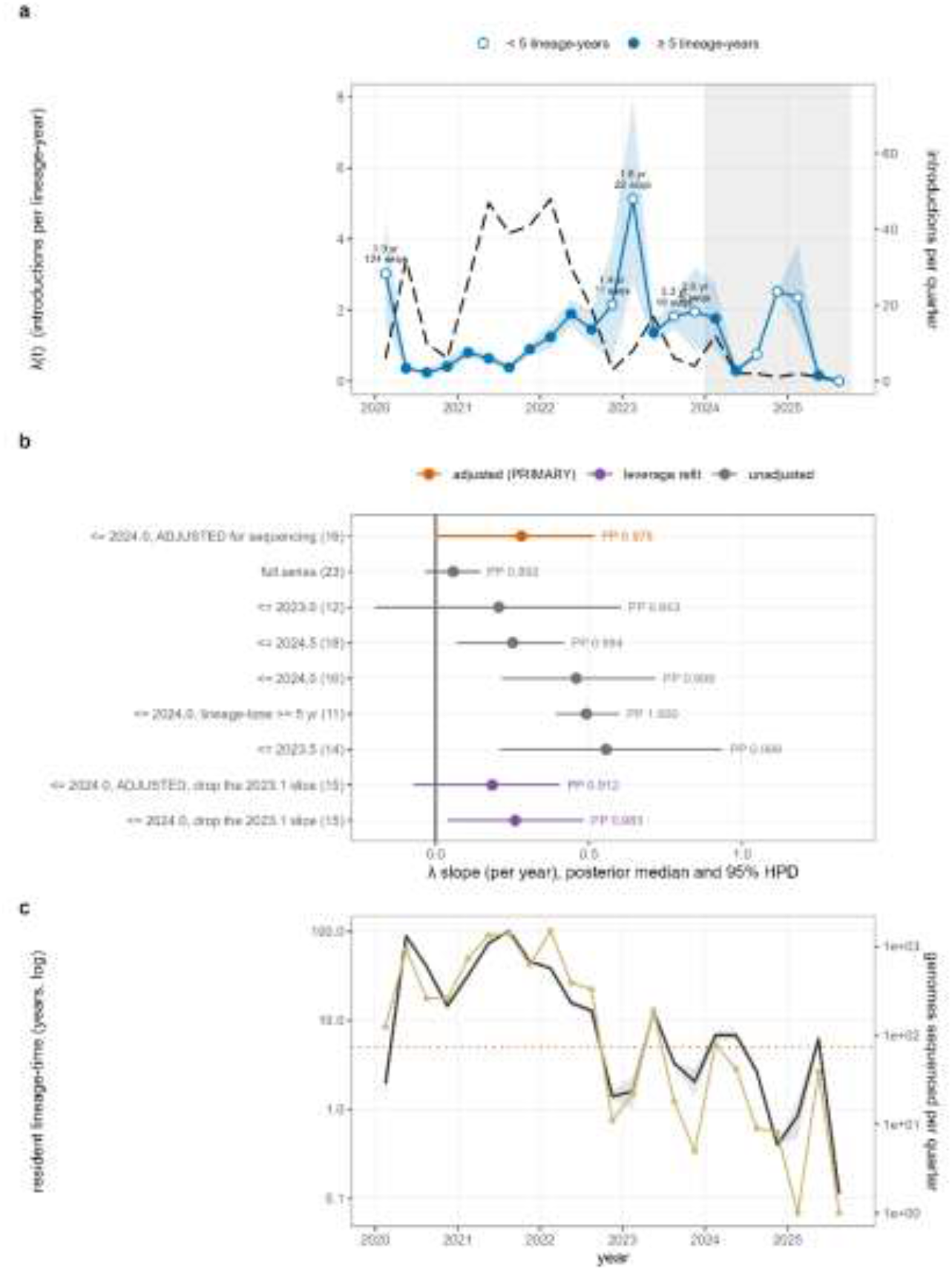
Changes in introductions per lineage year. (a) λ keeps rising after importation peaks. Blue = λ (median, 95% interval, 999 trees). Dashed grey = raw introductions per quarter, which decline. Hollow points (blue border) mark slices with under 5 lineage-years, where λ is unstable; the 2023.1 spike rests on 22 genomes. (b) The point estimate is positive in every window, but adjustment attenuates it, and the lower bound is unstable. Adjusting for sequences per slice reduces the estimate by 37% and brings the lower bound to 0.011. The λ 2023.0 window includes zero. PP = posterior probability. (c) The denominator tracks sequencing effort. Grey = resident lineage-time. Gold = genomes sequenced; log lineage-time rises with log sequences at slope 0.748 (95% HPD 0.715-0.785, R^2^ = 0.92), so λ is partly confounded with surveillance intensity. Dotted line = the 5-year floor.

That denominator is estimated from sampled genomes and scales with sequencing effort across quarterly slices, and log lineage-time rises with log genomes sequenced at slope 0.748 (95% HPD 0.715 to 0.785, R² = 0.92), so the two series track each other across two orders of magnitude (Fig. 4c). A positive λ trend is therefore equally consistent with sustained import dependence and with declining surveillance. Adjusting for sequencing gives 0.289 (0.012 to 0.524, PP 0.978) on the raw scale and 0.083 (−0.209 to 0.342, PP 0.720) on the logarithmic scale; excluding the single thinnest slice, which rests on 22 genomes, reduces the adjusted estimate to 0.148 (−0.105 to 0.371). It excludes zero under one of four specifications (Fig. 4b).

P_local trends negatively under every variant and threshold, and lineages introduced later do not persist longer (slope −0.025 per year, p = 0.48). Neither is independent of λ. All the functions of inferred lineage-time or lineage duration, so a decline in sequencing alone would move all three in the observed direction, and the persistence regression is additionally biased toward finding a decline because the end of the series truncates recently introduced lineages.

### 3.5 Bangladesh-enriched substitutions arise locally but rarely establish

Twenty-three substitutions were enriched in Bangladeshi genomes relative to a lineage- and month-matched global comparator (Table. S3). Twenty-two could be mapped to Bangladeshi lineages; ORF1a:H1500Y could not, as no occurrence was resolvable to a delineated lineage. Three further characters (S: A879S, ORF3a: V225F, ORF1a: S2224F) were excluded because their reconstructions failed to converge, and one (ORF1a: D981G) because all 92 of its carriers derive from a single sequencing laboratory whose overall share of Bangladeshi genomes is 5.2% (Table. S6). Here, 18 mutations, comprising 171 lineage–substitution occurrences across 124 of the 395 Bangladeshi transmission lineages (31%), were reported.

These 18 mutations are not 18 independent findings, as clustering substitutions at the overlap of their Bangladeshi carrier sets resolves them into 11 independent signals, three co-inherited haplotype blocks, and eight unlinked substitutions, and the count is identical at Jaccard thresholds of 0.30 and 0.50 (Table. S9). ORF1a: D983A and ORF3a: F28L are carried by exactly the same 75 genomes and are reported as one signal rather than two. Counts of distinct Bangladesh-enriched changes are given over signals below, and counts relating to the reconstruction itself are given over characters, and the two are distinguished throughout.

Origin classification was bimodal (Fig. 5a), and the highest root presence probability among locally arising occurrences was 0.109 and the lowest among imported occurrences 0.998; exactly one occurrence of 171 falls between them, ORF7a: L116F in BDTL-007, root-presence 0.663, carried by 126 of 181 genomes, and is reported as ambiguous. Any threshold in that interval yields identical classification. 117 occurrences were imported, and 53 arose locally, distributed across 16 of the 18 mutations and 10 of the 11 independent signals.

**Figure 5:**
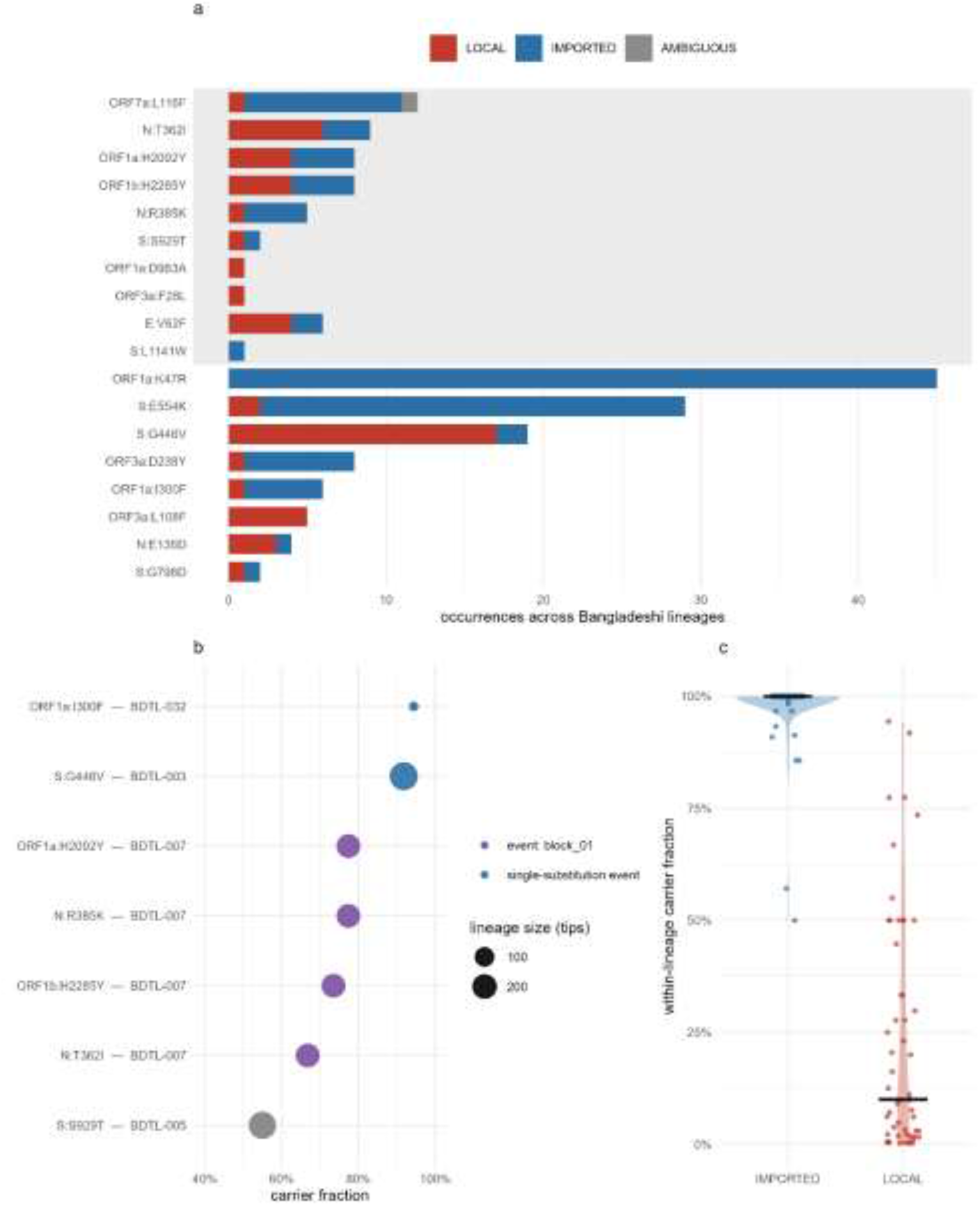
Locally enriched mutations in Bangladesh compared with global imports. (a) 18 substitutions, 171 occurrences resolving into 11 independent signals; classification threshold-insensitive (0.109 vs 0.998). Grey bands mark haplotype blocks. (b) Median within-lineage carrier fraction is 0.100 for locally arising occurrences; no locally arising occurrence reaches fixation (0/53). Imported occurrences are shown for context only: they are present at the lineage root by construction, so their carrier fraction is a property of the classification and is not a comparator. (c) Seven establishment occurrences in four lineages resolving into four independent events. The four BDTL-007 occurrences share a carrier set (haplotype block 1) and are counted once. One clade-wide sweep: S: G446V in BDTL-003 (BDTL_003; AY.131, 267/291 = 91.8%).

Two cases anchor the classification. ORF1a: K47R has zero local occurrences of 45, and it is imported in every lineage it enters and carried by every genome of each, a negative control confirming that lineage-defining substitutions are recognized as arriving with their lineage. Spike G446V is locally arising in 17 of 19 occurrences, is unlinked to any other enriched substitution, and has the highest local expansion rate of any substitution with three or more local occurrences and 11 of its 17 local occurrences (65%) achieved onward transmission, the profile expected of a change arising repeatedly within Bangladesh rather than travelling on a haplotype.

Local origin is common, but lineage-wide establishment is rare. Locally arising substitutions were carried by more than one genome of their host lineage in 27 of 53 occurrences (50.9%). This rate against imported occurrences was not compared, as an imported substitution is present at its lineage root by construction and is carried by more than one genome whenever its host lineage is not a singleton, so the imported rate recovers the non-singleton fraction of the lineage-size distribution (48.2%) rather than any property of the substitutions themselves. What they rarely achieve is lineage-wide frequency, and the median within-lineage carrier fraction of locally arising occurrences is 0.100 (IQR 0.022–0.447), and not one of the 53 reaches fixation (Fig. 5b).

Seven occurrences in four lineages met our establishment criterion of at least ten carriers and half the lineage (Fig. 5c). Four fall in a single lineage, BDTL-007, where N: R385K, ORF1a: H2092Y, and ORF1b: H2285Y co-occur in 142 genomes, and N: T362I is present in a nested subset of 115. These four, together with the ambiguous ORF7a: L116F, constitute one of the three haplotype blocks identified above, so their co-occurrence is established from carrier data rather than assumed. This reflects stepwise accumulation within one lineage, and it was counted as a single event. Four independent establishment events therefore occurred: that series in BDTL-007, S: S929T in BDTL-005, ORF1a: I300F in BDTL-032, and S: G446V in BDTL-003.

Only three of the seven establishment occurrences lie in lineages recovered in the posterior sample. All four BDTL-007 occurrences sit in a lineage whose exact tip set was never recovered across 999 trees (Table S4), so the composition of that event is uncorroborated even though its constituent substitutions are individually well resolved. The remaining three lie in lineages at posterior support 0.960 (BDTL-032), 0.964 (BDTL-003), and 0.754 (BDTL-005). One of the four also requires a distinction: ORF1a: I300F is present in all 850 Bangladeshi B.1.1.25 genomes and is effectively a lineage-defining substitution of that Pango lineage (Table S7); its establishment event is its local origin within BDTL-032, a separate B.1.1.25 lineage.

One locally arising substitution swept its lineage. Spike G446V arose within BDTL-003, an imported lineage of 291 genomes, 97.9% AY.131 (Delta, clade GK), posterior support 0.964, spanning 31 districts, TMRCA 2021.393, reached 267 of them, 91.8%, with a root-presence probability of 0.0009. Because BDTL-003 comprises almost all Bangladeshi AY.131 genomes, the sweep occurred at the level of the entire imported lineage rather than within a subclade of it: G446V is carried by 268 of 287 Bangladeshi AY.131 genomes (93.4%). Globally, AY.131 does not carry this substitution, which is why it is enriched in Bangladesh, and its absence from the lineage root establishes that it arose after the lineage was introduced.

Section 3.2 provides a mechanistic explanation for this asymmetry where the median lineage persists 62 days and half leave no sampled descendant, a substitution arising after establishment has little time to spread before its host lineage is lost from view. Because persistence is measured from the inferred TMRCA and contains a median detection lag of 46 days, that figure describes the interval over which lineages remain observed and bounds true persistence from below. Local mutation is neither rare nor disadvantaged in transmission, however, local fixation is rare because its host lineages remained observable for a short interval.

## 4 Discussion

Three hundred and sixty-one inferred introduction events of SARS-CoV-2 into Bangladesh are detectable between March 2020 and July 2025, of which slightly more than half leave a single sampled genome and no evidence of onward transmission. Introductions are concentrated in two corridors, India and the Gulf, which remain elevated after two independent controls for sampling bias, while three regions fall significantly below the controlled expectation. Within that imported diversity, 53 of 171 substitution occurrences arose after their host lineage was introduced, and just over half of these were carried by more than one genome; none reached fixation within its lineage. Four establishment events occurred, and one substitution, Spike G446V, swept an imported Delta lineage to 91.8%. Importation determined which lineages were available to circulate, local evolution determined what happened within them, and was rarely observed to complete.

### Import-driven lineage composition at low sequencing coverage

The introduction count, singleton fraction, and median persistence all depend on sequencing coverage and would shift with deeper sampling. They establish that importation was continuous and numerous, they do not establish that it exceeded local diversification by any particular factor, and the quantitative claim was not made. The values are comparative rather than absolute as they place Bangladesh in the same qualitative regime as settings sampled two orders of magnitude more deeply (Du Plessis et al., 2021; Lemey et al., 2021). In this study, 361 lineages are delineated from 4,598 genomes representing under one per cent of reported cases, and the size distribution is similarly heavy-tailed with a comparable singleton fraction. That the qualitative pattern survives a two-order-of-magnitude difference in coverage is itself informative, but it is not a quantitative equivalence and should not be read as one.

One line of evidence in this study is less coverage dependent than the counts, and it concerns the structure of variant diversity rather than its abundance. Nearly three quarters of Bangladeshi Beta genomes descend from a single sub-lineage attributed to Africa at posterior 0.975, in a country where Africa sits at the null model for source attribution. A single introduction accounting for nearly three-quarters of the sampled diversity of a variant is a founder signature that local diversification does not produce that concentration, whereas one arrival followed by expansion does (Gu et al., 2022). Because the numerator and denominator are drawn by the same sampling process, this proportion is less sensitive to absolute sequencing depth than an introduction count, though it remains sensitive to non-random sampling across time, geography, and outbreaks.

### Source attribution is consistent with labor-migration and air-travel corridors

The elevation of India and the Gulf countries and the depression of East Asia and South-East Asia is not dependent on proximity. Although Bangladesh borders India, it does not border the Gulf, and its nearest neighbors after India, namely South-east Asia, are precisely the regions that fall below the null model control. Instead, the pattern is consistent with the structure of Bangladeshi overseas employment and the direct air routes that serve it, although neither migration volumes nor flight data entered the model (Ahsan et al., 2020). East Asia is both the region in which the root is reconstructed, at posterior 0.9994, and the region with the largest deficit, that is, a pattern consistent with an early seeding route that did not sustain later introductions, though a root state reflects the most probable ancestral assignment under this model and sampling scheme rather than a documented epidemiological corridor. The settings analyzed in the comparable literature do not include a large population employed overseas in a single region, so we are not aware of a directly comparable corridor.

India and the Gulf were reported as jointly dominant rather than ranked. The primary analysis separates them (difference-in-differences 2.66 pp, p = 0.0098), but the ordering reverses under the uncontrolled complete data null estimates.

### Local evolution is not suppressed; it is rarely observed to complete

As per the lineage dynamics, the median lineage is observed across roughly two months, which includes the detection lag and therefore is a lower bound, and half left no sampled descendant. A substitution arising after establishment consequently has little opportunity to spread before its host lineage leaves the record. This same mechanism underlies the observation, in more densely sampled settings, that most within-host and within-lineage diversity fails to propagate between transmission chains (Lythgoe et al., 2021), but here it operates at the level of whole transmission lineages rather than individual hosts.

Substitutions carried on a shared genomic background are not independent observations, and clustering the eighteen reported substitutions on the overlap of their carrier sets resolves them into eleven independent signals, three co-inherited blocks and eight unlinked substitutions, a count stable across the range of clustering thresholds examined. Two of the characters share an identical carrier set and are one change reported twice. This matters for the establishment count as well as for the enrichment test, four of the seven establishment occurrences fall in a single lineage and share a carrier set, so they represent one stepwise accumulation rather than four independent events.

Spike G446V is the exception. It arose independently in 17 of 19 host lineages, is one of the eight unlinked signals, and has the highest local expansion rate of any character with three or more local occurrences. It is absent from the root of BDTL-003, an imported AY.131 lineage, at posterior probability 0.0009, and is carried by 267 of its 291 genomes. Residue 446 lies within the class 3 antibody epitope (Barnes et al., 2020), and substitutions at this position contribute to escape in other genetic backgrounds (Cao et al., 2022; Greaney et al., 2022). The substitution reported here is a different one at the same position, arising in a Delta background more than a year before BA.1 circulated in Bangladesh, and do not claim it confers the phenotype characterized for G446S. What is notable is that repeated independent origin at an antibody-contact residue within one country is compatible with local selection, but the present design cannot separate selection from founder effects, superspreading, linkage, or drift (Focosi et al., 2023; Van Dorp et al., 2020).

The expansion of G446V within BDTL-003 is equally consistent with a founder effect followed by lineage-level expansion for reasons unrelated to the substitution, and because BDTL-003 comprises almost all Bangladeshi AY.131 genomes, the sweep occurred at the level of the entire imported lineage rather than within a subclade. The data show that the substitution arose after the lineage was introduced, recurred, and reached clade-wide frequency once.

The denominator of importation per unit resident lineage-time is not observed but estimated from sampled genomes, and across quarterly slices log lineage-time rises with log genomes sequenced at slope 0.748 (R² = 0.92). A positive slope in λ is therefore consistent both with sustained import dependence and with a measurement artefact of declining surveillance, and this dataset cannot separate them. Sequencing entered the adjustment on the raw scale, giving 0.289 per year (95% HPD 0.012 to 0.524; PP 0.978); entering it on the logarithmic scale on which the confounding was itself characterized gives 0.083 (−0.209 to 0.342; PP 0.720). The highest λ estimate within the analysis window rests on approximately 22 sequenced genomes and 1.6 lineage-years; excluding that single slice reduces the unadjusted slope from 0.459 to 0.260 and the adjusted slope from 0.289 to 0.148 (−0.105 to 0.371; PP 0.876). A post-hoc check on an observed quantity, whether lineages introduced later persist longer, finds no increase (slope −0.025 per year, p = 0.48), and P_local trends negatively under every variant and threshold, though its interval always includes zero. All three statistics point in a direction consistent with continued import dependence, but they are not independent evidence. Each is a function of resident lineage-time, which is inferred from the same genomes that generate the introduction counts, so a decline in sequencing alone would shorten observed persistence, reduce inferred lineage-time, raise λ and lower P_local simultaneously. Their concordance is therefore expected under a sampling constraint. The problem is general as any rate whose denominator is inferred from the sampling process that generates its numerator will track surveillance intensity rather than epidemiology. Unequal sampling is already known to distort phylogeographic reconstruction (Liu et al., 2022) and inferred importation counts to depend on sampling strategy even under systematic high-coverage surveillance (Goliaei et al., 2024), where sequencing effort falls by more than an order of magnitude, as it did here, that dependence dominates.

Twenty of the 53 named lineages, including four of the ten largest, have tip sets that never recur across the posterior sample. The correct reading is not that these clusters may not exist but that their exact membership is not determined. This bears most on BDTL-007, which carries four of the seven establishment occurrences; that event should therefore be regarded as uncertain, leaving three posterior-corroborated establishment events.

Recombination violates the single-tree assumption, so their inferred divergence times are less reliable; these were retained to preserve tip-set correspondence between the maximum clade credibility tree and the posterior sample. Furthermore, seven background genomes are not human transmission samples: four environmental, one canine, and two mouse-adapted laboratory strains whose substitutions were acquired during serial passage (Leist et al., 2020). All seven fall in Rest-of-World, and excluding them shifts no regional background share by more than 0.10 pp.

Submitting laboratory records were obtained for every Bangladeshi genome, so the screen is not limited by coverage. Five substitutions are nonetheless fixed within their Pango background and therefore have almost no carriers in a stratum containing a non-carrier; their permutation p-values are the absence of a test rather than evidence of no clustering. All five are lineage-defining, and a substitution present in 100% of a lineage across many laboratories cannot be a protocol signature, since batch effects are laboratory-associated and therefore variable within a lineage. A sixth, ORF3a: D238Y, is significant under the coarser stratification and loses both significance and more than half its permutable carriers under the finer one; it was reported it as inconclusive rather than cleared. The block structure has an implication. When enrichment is used to select candidates for downstream analysis, cluster candidates by shared carriage before counting. Eighteen substitutions here are eleven changes, and two of them are one; a study reporting the larger number would have overstated independent local evolution by nearly two thirds (Table S8).

Introductions were frequent and continuous across 2020–2025, and most left no sampled descendant. India and the Gulf supplied disproportionately more of them than sampling alone would produce, a pattern consistent with labor-migration and air-travel structure rather than with geographical proximity. Whether dependence on importation changed over time could not be determined, because the relevant statistic has a denominator inferred from the same sampling process that generates its numerator. Roughly a third of enriched-substitution occurrences arose after their host lineage entered the country, so local evolution was common rather than exceptional. And one locally arising substitution, Spike G446V, reached clade-wide frequency, arising independently in most lineages carrying it and sweeping an imported Delta lineage to 91.8%. Importation determined which lineages were available to circulate in Bangladesh; local evolution determined what happened within them, and was rarely observed to complete before its host lineage left the genomic record.

The wider point is methodological. The qualitative signature of import-driven lineage composition survives at sequencing coverage two orders of magnitude below the settings in which it was established, while the quantitative statistics built on that signature do not. That distinction is not specific to Bangladesh. It will recur wherever surveillance intensity varies by an order of magnitude across a study period, which is to say across most of the world.

## Ethics statement

This study used only SARS-CoV-2 genome sequences and associated metadata that were publicly deposited in the GISAID EpiCoV database by the originating and submitting laboratories. No human participants were recruited, no biological specimens were collected, and no identifiable personal information was accessed at any stage. Ethical approval was therefore not required for this analysis.

This research received no specific grant from any funding agency

## Author contributions

Sabik Khair - Conceptualization, Data curation, Analysis, Methodology, Software, Visualization, Writing; Mst. Noorjahan Begum - Analysis, Data curation, Review and Editing; Yeasir Karim - Methodology and Writing

## Data availability

All genomes analyzed are available from GISAID EpiCoV under EPI_SET_260813zw (https://doi.org/10.55876/gis8.260813zw). We acknowledge the contributing, originating, and submitting laboratories in Table S9. We deposited the analysis code, trait files, delineation and attribution outputs, and the scripts generating every figure and table at Zenodo (https://zenodo.org/uploads/22684629), along with a table mapping every number reported in this paper to the file and script that produced it.

