## Supplementary figures and images for "Repeated SARS-CoV-2 Introductions with Limited Local Establishment in Bangladesh under Genomic Surveillance, 2020–2025"

### Fig. S1

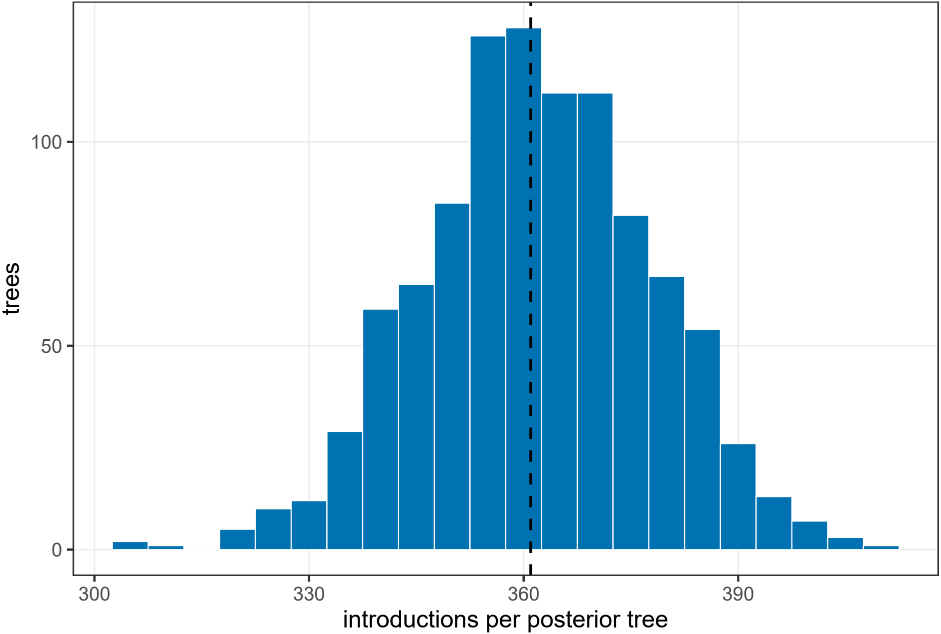

### Fig. S2

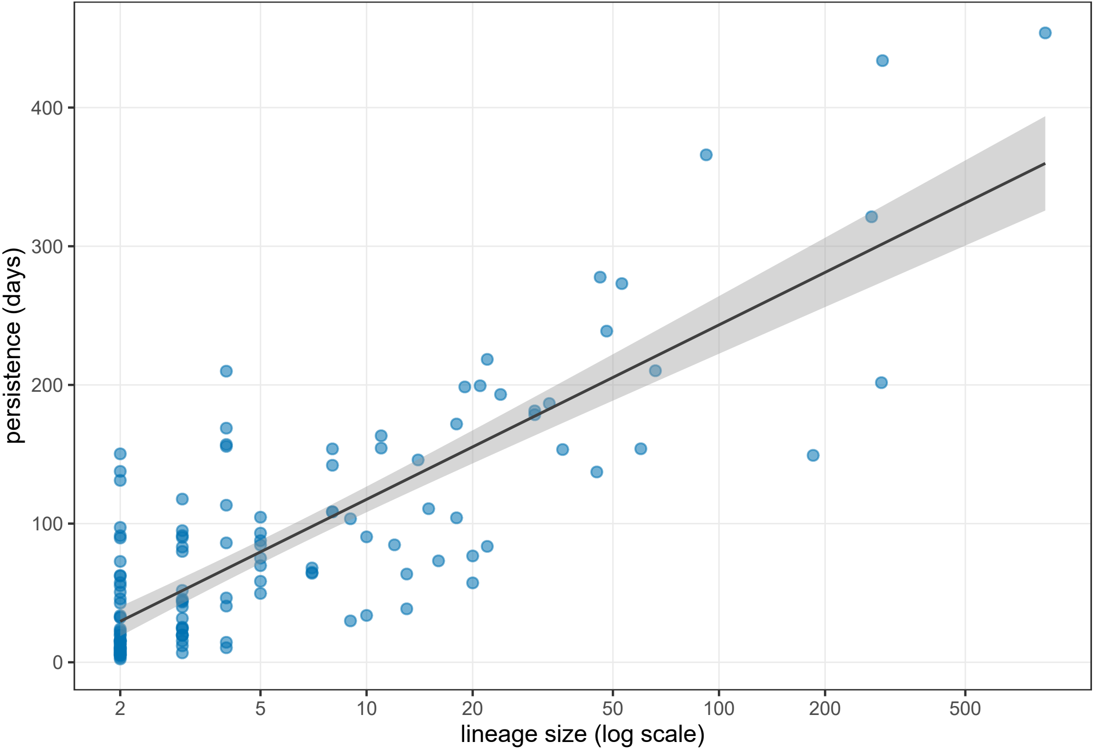

### Fig. S3

**Figure S3**

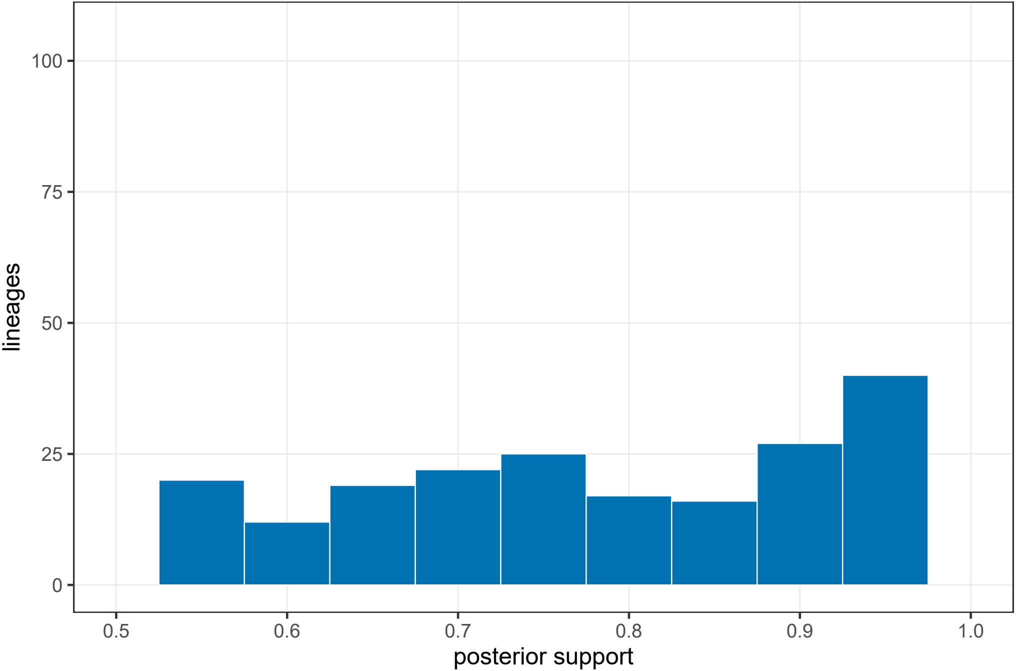

### Fig. S4

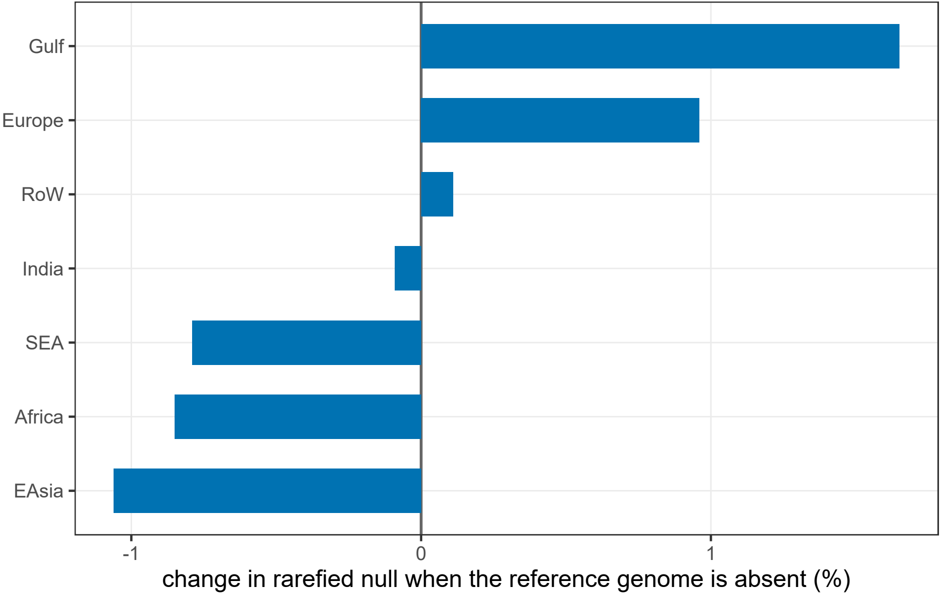
